# Anxiety, depression, and insomnia among nurses during the full liberalization of COVID-19: A multicenter cross-sectional analysis of the high-income region in China

**DOI:** 10.1101/2023.03.09.23286785

**Authors:** Julan Xiao, Lili Liu, Yueming Peng, Yi Wen, Xia Lv, Lijun Liang, Yi Fan, Jie Chen, Yanru Chen, Hongying Hu, Weisi Peng, Haiyan Wang, Weixiang Luo

**Affiliations:** Department of Thoracic Surgery, Shenzhen People’s Hospital (The Second Clinical Medical College, Jinan University; The First Affiliated Hospital, Southern University of Science and Technology), Shenzhen 518020, Guangdong, China; Shenzhen Clinical Research Centre for Geriatrics, Shenzhen People’s Hospital, Shenzhen 518020, Guangdong, China; Department of Psychology, School of Public Health, Southern Medical University, Guangzhou, Guangdong, China; Department of Nursing,Shenzhen People’s Hospital (The Second Clinical Medical College,Jinan University;The First Affiliated Hospital,Southern University of Science and Technology),Shenzhen 518020,Guangdong,China; Department of Nursing,Longhua Branch of Shenzhen People’s Hospital (The Second Clinical Medical College,Jinan University;The First Affiliated Hospital,Southern University of Science and Technology),Shenzhen 518020,Guangdong, China; Department of Cardiovascular Medicine,Shenzhen People’s Hospital (The Second Clinical Medical College,Jinan University;The First Affiliated Hospital,Southern University of Science and Technology),Shenzhen 518020,Guangdong,China.; Department of Respiratory and Critical Care Medicine,Shenzhen People’s Hospital (The Second Clinical Medical College,Jinan University;The First Affiliated Hospital,Southern University of Science and Technology),Shenzhen 518020,Guangdong,China

**Keywords:** Anxiety, depressive symptoms, insomnia, perceived stress, nurses, epidemiology, full liberalization of COVID-19

## Abstract

**Aims and objectives:** This study demonstrates the impact of the full liberalization of COVID-19 on the psychological issues and the prevalence rate and associated factors of depressive symptoms, anxiety, and insomnia among frontline nurses.

**Background:** It has been demonstrated that frontline nurses fighting against the epidemic were under great psychological stress. However, there is a lack of studies assessing the prevalence rates of anxiety, depression, and insomnia among frontline nurses after the full liberalization of COVID-19 in China.

**Design:** Cross-sectional study.

**Methods:** Of 1766 frontline nurses were invited to complete a self-reported online questionnaire by convenience sampling. The survey included six main sections: the Patient Health Questionnaire, the Generalized Anxiety Disorder Scale, the Insomnia Severity Index, the Perceived Stress Scale, sociodemographic information, and work information. Multiple logistic regression analyses were applied to identify the potential risk factors for psychological issues. Reporting of this research according to the STROBE checklist.

**Results:** 90.83% of frontline nurses were infected with COVID-19, and 33.64% had to work while infected COVID-19. The overall prevalence of depressive symptoms, anxiety and insomnia among frontline nurses was 69.20%, 62.51%, and 76.78%, respectively. Multiple logistic analyses revealed that job satisfaction, attitude toward the current pandemic management, and perceived stress were associated with depressive symptoms, anxiety, and insomnia.

**Conclusions:** This study demonstrated that the full liberalization of COVID-19 had a significant psychological impact on frontline nurses. Early detection of mental health issues and preventive and promotive interventions should be implemented according to the associated factors to improve mental health of nurses.

**Relevance to clinical practice:** This study highlighted that nurses were suffering from varying degrees of depressive symptoms, anxiety, and insomnia, which needed early screening and preventive and promotive interventions for preventing a more serious psychological impact on frontline nurses.

**Patient or Public Contribution:** No Patient or Public Contribution.

## 1 INTRODUCTION

Coronavirus disease 2019 (COVID-19) is a rapidly spreading disease with high infectivity and high morbidity and mortality rates worldwide caused by severe acute respiratory syndrome-novel coronavirus-2 (SARS-CoV-2) (Jakovljevic et al., 2020). The pandemic has caused a global threat to health and life. By the end of April 2020, approximately 90,000 health care workers worldwide were infected, and more than 600 nurses died (Dawn News, 2020). As such, nurses who care for COVID-19 patients are at a high risk of infection. To combat COVID-19, in affected countries such as China, measures have been implemented to prevent the spread of the virus, such as quarantine, daily nucleic acid testing, temperature screening and screening of patients with COVID-19, and setting up fever clinics and isolation units in many hospitals (National Health Commission of the People’s Republic of China, 2023). On December 27, 2022, the National Health Commission of the People’s Republic of China issued a circular on further optimizing prevention and control measures against COVID-19 and suddenly adopted a policy of full liberalization, called daily nucleic acid testing or temperature screening of patients with COVID-19 infection. In a very short time, most people, whether they were medical staff or the general population, were infected. There was a lack of preparation for defeating COVID-19 due to sudden full liberalization, and nurses faced challenges and difficulties such as the huge increase in the number of critically ill COVID-19 patients. Many nurses were infected and had to work while ill, and nursing work stress increased. Nurses were physically and psychologically exhausted from intensive, heavy workloads and long-term usage of personal protective equipment (Liu et al., 2020; Doo et al., 2020).

## 2 BACKGROUND

Nurses consitute the largest proportion of hospital health care professionals (United States Bureau of Labor Statistics, 2023), spend more time caring for patients than other health care workers, and play an essential role in the treatment, care, and control of patients during disease outbreaks, disasters, and emergency situations (Doo et al., 2020; Labrague & de Los Santos, 2021) . In addition, nurses have the closest proximity to COVID-19 patients worldwide. Previous research has shown that frontline health care providers have a high risk of mental health conditions during the COVID-19 pandemic, mainly fear, anxiety, depression, and insomnia. Approximately 91.2% of 1837 nurses working in Wuhan, China, showed fear of infection and death as well as fear of nosocomial spreading to their loved ones (Hu et al., 2020). A systematic review and meta-analysis of global health care professionals showed that the pooled prevalence of psychological problems for anxiety, depression, and distress was 40%, 37%, and 37%, respectively (Saragih et al., 2021). A cross-sectional study reported that the prevalence of anxiety, depression, insomnia, and nonspecific distress symptoms in Chinese frontline health care workers was 44.6%, 50.4%, 34.0%, and 71.5%, respectively (Lai et al., 2020). Therefore, it is necessary to pay special attention to the psychological health of frontline nurses.

Nurses were at the forefront of defeating the pandemic and were therefore at risk of developing high levels of stress and physical and psychological depletion. They were affected by a variety of stressors in their workplaces because they were confronted with a complex psychological conflict between their responsibility, dedication, and mission as nurses to provide health and care for patients with a very dangerous virus and their right to protect themselves from the virus (Hu et al., 2020; Kim, 2018). After the sudden full liberalization of COVID-19, nurses experienced acute and exacerbated stress (e.g., more COVID-19 patients, direct exposure to diseases, longer workload and work shifts, and high rates of infection by COVID-19), whereas there was a shortage of resources (e.g., a lack of protective equipment, insufficient facilities, insufficient nursing staff, lack of training or experience in caring for seriously ill COVID-19 patients) (Lorente, Vera & Peiró, 2021). In a systematic review of the psychology of frontline health care staff caring for COVID-19 patients, the prevalence of stress was 45% (Salari et al., 2020). Stress is the most common issue and an integral part of human life. Hospital staff in charge of admitting and directly caring for COVID-19 patients were subjected to pressure that adversely impacted their health and job satisfaction (Salari et al., 2020). Stress can increase anxiety and depression, jeopardize physical, mental, and social health, and even lead to suicidal thoughts (Kapur et al., 2016). More importantly, poor psychological health among nurses not only has been linked to negative outcomes such as psychological distress, depression, anxiety, and burnout but also hinders professional performance, in turn leading to the deterioration of the quality of patients’ care (Lorente, Vera & Peiró, 2021). Thus, frontline nurses should maintain excellent physical and psychological health.

Nurses working during the global pandemic were physically and psychologically exhausted and experienced severe anxiety, depression, and stress. However, there is a lack of studies exploring anxiety, depression, perceived stress, and sleep disorders among nurses during the sudden full liberalization of COVID-19 in China. In particular, most medical staff were also infected during this specific period. In addition to psychological health problems, some physical symptoms and manifestations of the virus, such as pain, fatigue, cough, expectoration, sore throat, and weakness, caused additional workload to health care systems.

This study aimed to determine the prevalence rates of anxiety, depression, and insomnia among frontline nurses during the full liberalization of COVID-19 in Shenzhen, China, and to explore risk factors and protective measures to provide a theoretical basis for the early identification and intervention of negative emotions for these professionals. We hypothesize that perceived stress, job satisfaction, turnover intention, attitude toward current pandemic management, and perceived health status are associated with the risk of depression, anxiety, and insomnia. To our knowledge, this is the first study to determine the prevalence of mental health issues after the full liberalization of COVID-19. Consequently, our study will make a positive contribution to the literature on organizing preventive mental health services and help plan emergency responses to future infectious disease outbreaks.

## 3 METHOD

### 3.1 Study Design, Setting, and Participants

This was a multicenter cross-sectional study conducted from December 27, 2022, to January 7, 2023, in six tertiary hospitals in Shenzhen. Convenience sampling was used for the recruitment strategy, and 1833 nurses volunteered to participate. The online questionnaire was shared via “Wenjuan Xing”, a professional questionnaire survey platform that is widely used in China (Zheng et al., 2021). The online survey was first disseminated through an instant messaging system, the WeChat group, to nursing managers at Shenzhen hospitals, who were encouraged to pass it on to other frontline nurses at Shenzhen hospitals. The inclusion criteria for participants were as follows: (1) nurses aged 18 and above; (2) normal language expression and understanding and the ability to understand the investigation content and cooperate with the research; and (3) the ability to give voluntary informed consent. The exclusion criteria were (1) history of mental illness and (2) serious physical diseases. Screening criteria included a response time of fewer than 200 seconds (Trauzettel-Klosinski, Dietz & IReST Study Group, 2012) to complete the survey and the deletion of incomplete or repeated answers. While reviewing the participants’ data, sixty-seven nurses completed the questionnaire in less than 200s or had missing data. In total, 1766 frontline nurses completed the questionnaire (response rate of 96.34%).

### 3.2 Procedures

This study was conducted in Shenzhen, which is one of the highest-income cities in China. All investigators received unified training on the questionnaire survey. The unified guidance on the questionnaire explained the purpose and completion method of this research. Each question was a mandatory item and could only be answered once by the same IP address. In addition, the nurses were told that the survey could be stopped at any time, and anonymity would be guaranteed. The online questionnaire began with informed consent. Frontline nurses needed to read the informed consent and chose the “agree” option to begin filling out the questionnaire; otherwise, the questionnaire could not be completed. To test the feasibility and suitability of the questionnaire, online pilot test including 15 frontline nurses was conducted. The nurses who participated were also asked for advice on questionnaire modification. Pilot test data were not used for the final statistics. The trained authors of this article distributed the final version of the questionnaires to the frontline nurses for data collection.

### 3.3 Variables and measurement

#### 3.3.1 Demographic characteristics

A general information questionnaire (created by the authors in line with the literature) included two parts. The first part collected data on the participants’ gender, age, educational background, working years, marital status, income per year, staff type, professional technical titles, night shifts per month, hospital type, and hospital department. The second part included job satisfaction, turnover intention, COVID-19 vaccination status, infection with COVID-19, working status, and compared with the outbreak period, the attitude toward the current epidemic.

#### 3.3.2 Perceived Stress

The 10-item Perceived Stress Scale (PSS-10) is based on the theory of psychological stress and was developed in 1983 by Cohen et al (Cohen, Kamarck & Mermelstein, 1983). The PSS-10 is used to measure how stressful an individual perceives events in daily life. The scale has previously shown good reliability and validity (Cohen, Kamarck & Mermelstein, 1983). The PSS-10 is a 10-item self-report scale; six items measure crisis perception and four items measure coping ability. Each item is rated on a 5-point Likert-type scale ranging from 0 indicating “never” to 4 indicating “very often”. The total score on the PSS-10 ranges from 0-40. Higher scores indicate that the individual’s perceived stress level is high. In the current study, the Cronbach’s alpha for the PSS-10 was 0.72, the Guttman Split-Half Coefficient was 0.88.

#### 3.3.3 Depressive symptoms

The severity of depression was assessed using the 9-item Patient Health Questionnaire (PHQ-9), which is a self-report validated screening instrument for use among patients and general populations. The questionnaire is based on the nine symptoms of depression in the US Diagnostic Standard for Mental Illness (DSM-IV) (Spitzer, Kroenke & Williams, 1999). Participants rate their experiences in the past 2 weeks, scoring each item from 0 to 3 (0=not at all, 1=several days, 2=more than half the days, and 3=nearly every day). Total scores on the PHQ-9 range from 0 (not at all) to 27 (extremely severe depression) (Kroenke, Spitzer & Williams, 2001) , the higher the score, the more obvious the state of depression. Regarding severity, PHQ-9 comprises five categories as follows: minimal/no depression (0-4), mild depression (5-9), moderate depression (10-14), or severe depression (15-21) (Kocalevent, Hinz & Brähler, 2013). In our study, the Cronbach’s alpha for the PHQ-9 was 0.93.

#### 3.3.4 Anxiety

We used the 7-item Generalized Anxiety Disorder (GAD-7) to evaluate the severity of anxiety, which is a self-rated scale with good reliability and validity (Kang et al; 2020). Each item was scored based on a 4-point Likert scale, where 0 and 3 points indicated ‘never’ and ‘almost every day’, respectively. Higher scores indicated higher exposure to anxiety. The total scores are categorized as follows: minimal/no anxiety (0-4), mild anxiety (5-9), moderate anxiety (10-14), or severe anxiety (15-21). Cronbach’s alpha in the present study was 0.97.

#### 3.3.5 Insomnia

The 7-item Insomnia Severity Index (ISI) was used to evaluate the severity of insomnia in the past week. The ISI is a measure of insomnia severity that has been shown to be valid and reliable. The total scores are categorized as follows: normal (0-7), subthreshold (8-14), moderate insomnia (15-21), or severe insomnia (22-28) (Morin, Belleville, Bélanger & Ivers, 2011). Cronbach’s alpha of the scale was 0.93 in this study.

### 3.6 Statistical analysis

Data were analyzed with SPSS version 26.0. Descriptive statistics, including frequencies and central tendencies, were calculated to characterize the sample’s demographic profile. Binary and multiple logistic regression analyses were performed to explore the potential factors influencing depression, anxiety and insomnia after the sudden full liberalization of COVID-19. Perceived stress was divided into four groups using quartiles, indicating the position of the score in the sample. The first quartile was 0-25%, the second was 25%-50%, the third was 50%-75%, and the fourth was 75%-100%. Odds ratios (ORs) and 95% confidence intervals (95% CIs) were obtained from the logistic regression models. *P*<0.05 was considered statistically significant.

### 3.7 Ethical considerations

This study was in line with ethical principles, and the contents of the questionnaire didn’t involve private and sensitive topics such as names. More importantly, the research was approved by the ethics committee of China People’s Hospital (Approval NO. LL-KY-2022004-01). All samples were given information about introducing the study and notified about their own right to withdraw at any time, and informed consent was sought from all eligible participants, which illustrated that they had understood the study in its entirety.

## 4 RESULTS

### 4.1 Sociodemographic and characteristics of the participants

The demographic characteristics of the 1766 participants are summarized in Table 1. Most of the participants (n=1694, 95.92%) were female, and the majority (n=1342, 75.99%) were younger than 40 years of age. A total of 30.81% (n=544) of respondents had less than 5 years of work experience, while 45.36% (n=801) reported working in hospitals for more than 10 years. A total of 81.99% (n=1448) of the sample were employed as contract staff with a primary title (n=1092, 61.83%). More than half of the nurses (n=1000, 56.63%) were married.

**Table1.**
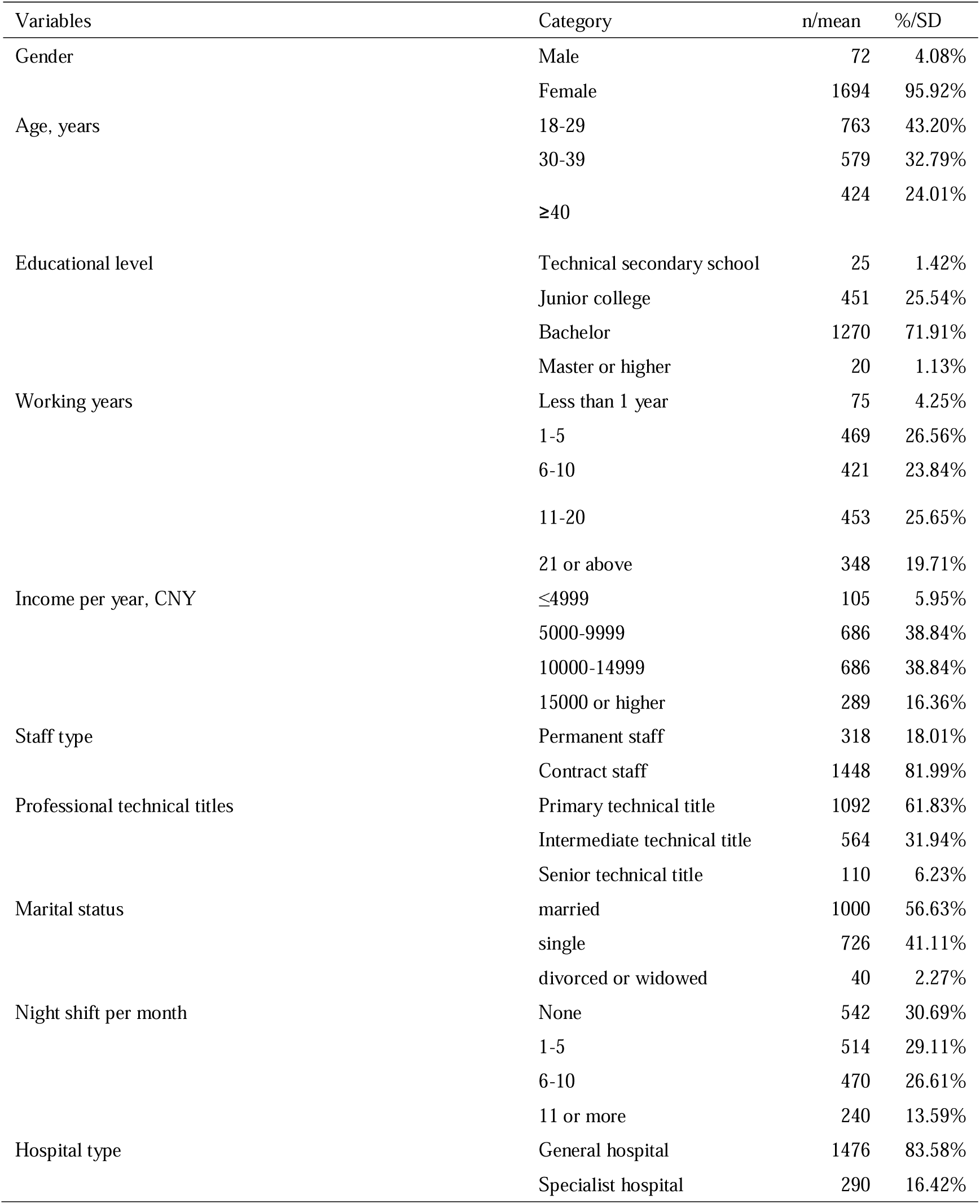
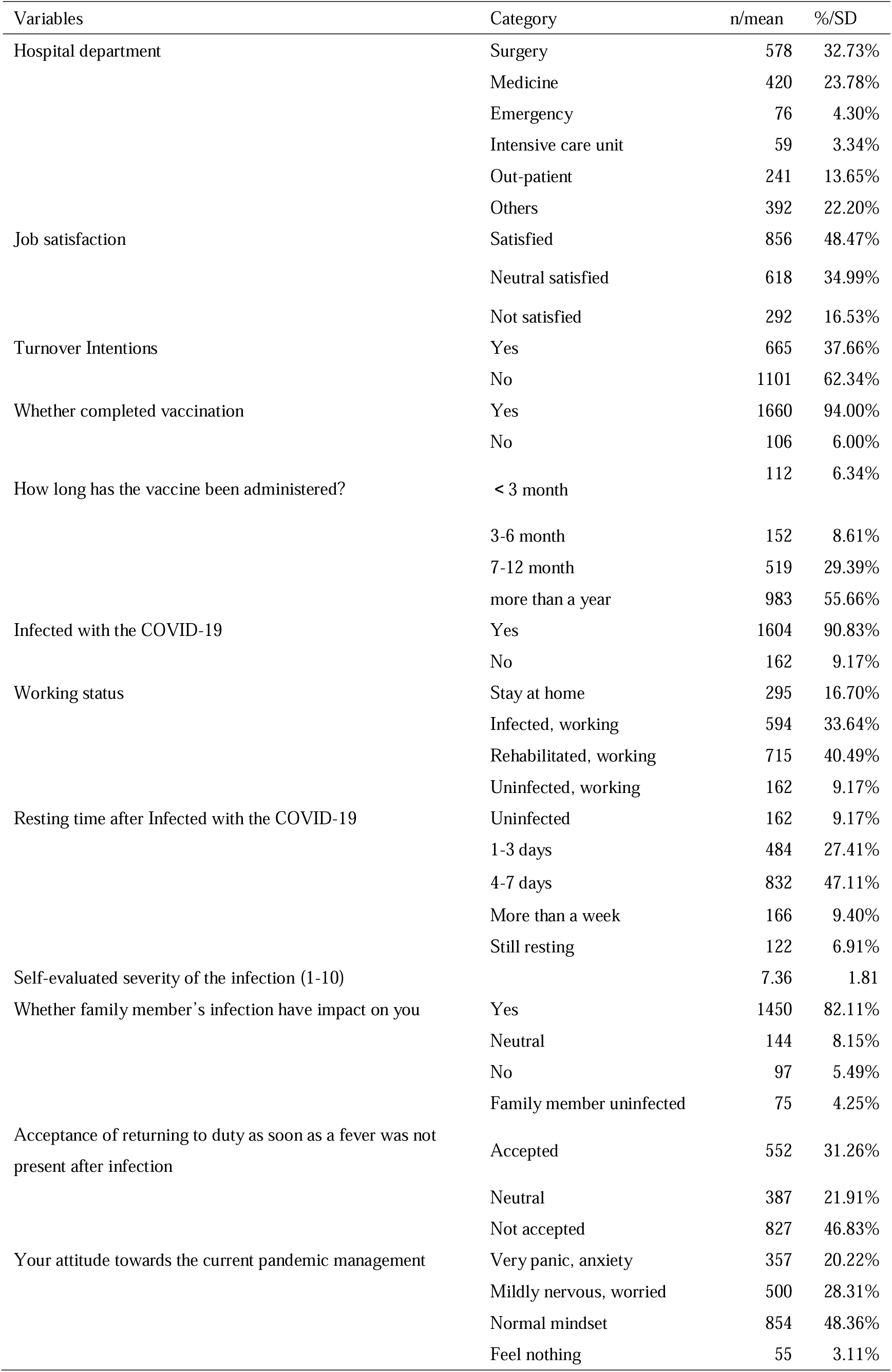
Demographic characteristics of the study subjects (n = 1766).

Regarding the characteristics of the nurses’ working situations, 30.69% (n=542) of nurses had no night shifts, while 13.59% (n=240) had 11 or more night shifts per month. The majority (n=1476, 83.58%) stated that they worked in general hospitals. Among the frontline nurses, 4.3% (n=76) worked in the emergency department and 3.34% (n=59) in the intensive care unit (ICU); 48.47% (n=856) were satisfied with their current nursing work, and only 16.54% (n=292) were not satisfied with their work. Furthermore, 37.66% (n=665) had turnover intentions. Overall, 90.83% (n=1604) were infected with COVID-19, and 33.64% (n=594) had to work while infected COVID-19, while 68.74% (n=1214) did not accept their return to duty as soon as a fever was not present after infection.

### 4.2 Prevalence of depressive symptoms, anxiety, and insomnia

The prevalence rates of depression, anxiety, and insomnia in the survey sample are shown in Table 2. The overall prevalence of depressive symptoms, anxiety and insomnia among frontline nurses was 69.20%, 62.51%, and 76.78%, respectively. Specifically, the participants indicated mild (n=538, 30.46%), moderate (n=352, 19.93%), and severe (n=332, 18.80%) depressive symptoms. Similarly, the participants reported mild (n=637, 36.07%), moderate (n=143, 8.10%), and severe (n=324, 18.35%) anxiety. The majority of the nurses reported mild (n=745, 42.19%) and moderate (n= 443, 25.08%) insomnia (Fig.1).

**Table2.** Prevalence of depressive symptoms, anxiety and insomnia among nurses during the early open up phase of the COVID-19 pandemic.

|  | Depressive symptoms |  | Anxiety |  | insomnia |  |
| --- | --- | --- | --- | --- | --- | --- |
|  | n | % | n | % | n | % |
| Absence of symptoms | 544 | 30.80% | 662 | 37.49% | 410 | 23.22% |
| Mild | 538 | 30.46% | 637 | 36.07% | 745 | 42.19% |
| Moderate | 352 | 19.93% | 143 | 8.10% | 443 | 25.08% |
| Severe | 332 | 18.80% | 324 | 18.35% | 168 | 9.51% |

**Fig.1.**
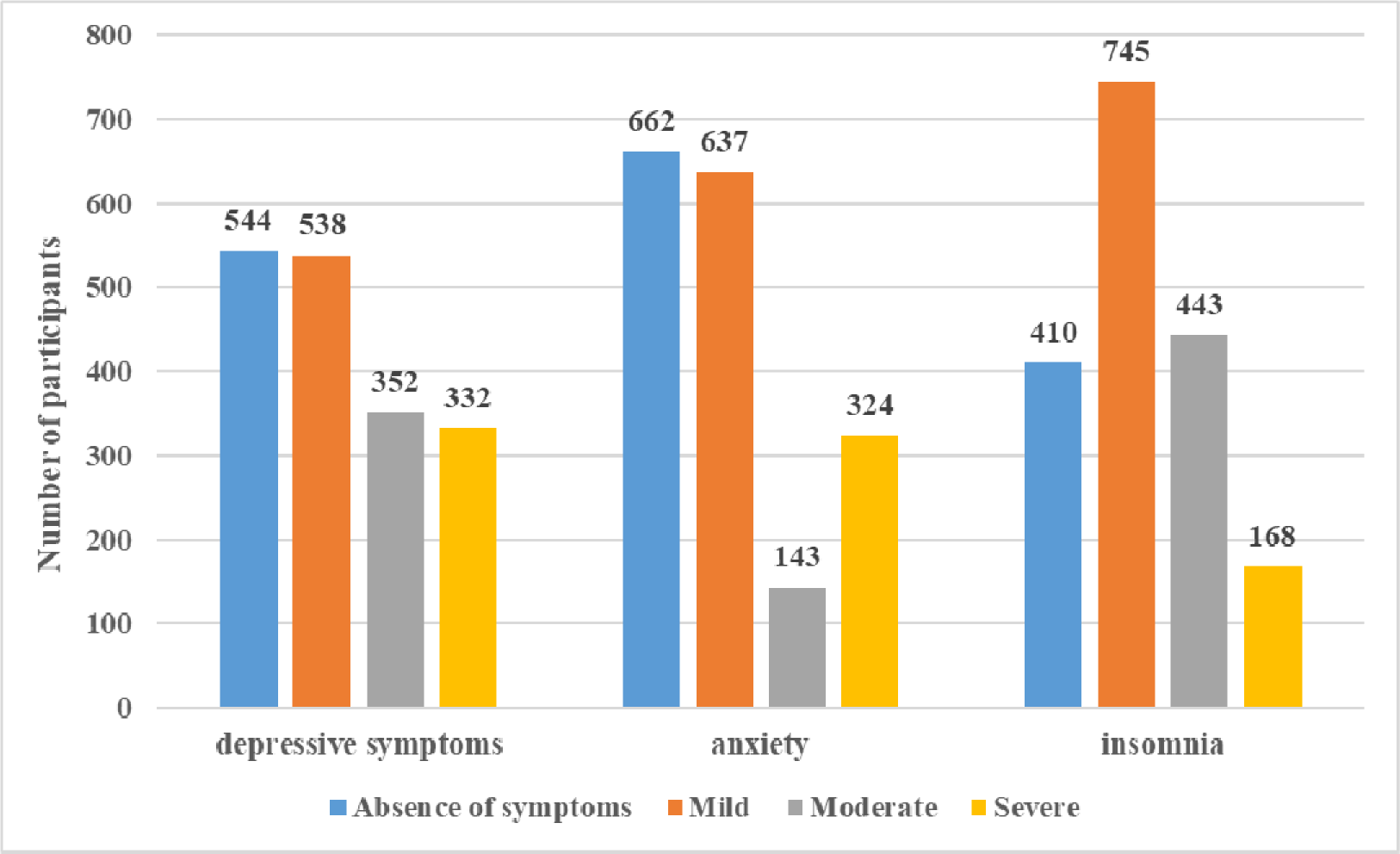
Number of participants experiencing depressive symptom, anxiety and insomnia (N=1766).

### 4.3 Risk factors associated with depressive symptoms

The results from the binary logistic regression analysis are presented in Table 3. Nurses with a primary technical title and intermediate nurses had a higher level of depression than those with a senior title. Nurses who were single (*OR*, 1.58; 95% *CI*, 1.12-2.22), had 11 or more night shifts per month (*OR*, 1.70; 95% *CI*, 1.08-2.67), had turnover intentions (*OR*, 1.69; 95% *CI*, 1.29-2.21) were at greater risk of depression. Nurses were dissatisfied or had neutral satisfaction with their current nursing work were at greater risk of depression than those who were satisfied. A negative attitude toward the current pandemic management and nurses with higher total PSS-10 scores also had a higher risk of depression.

**Table3.**
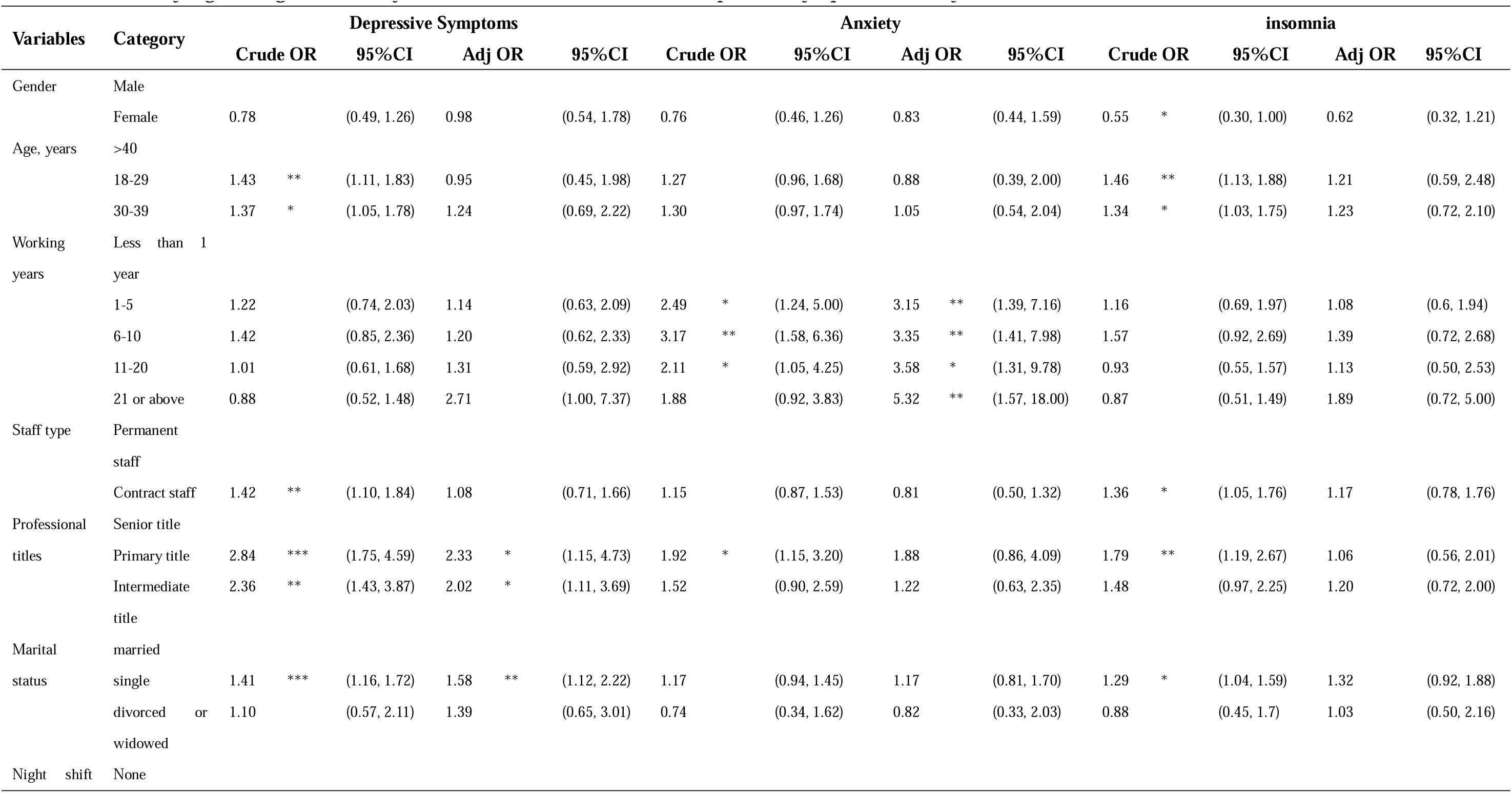
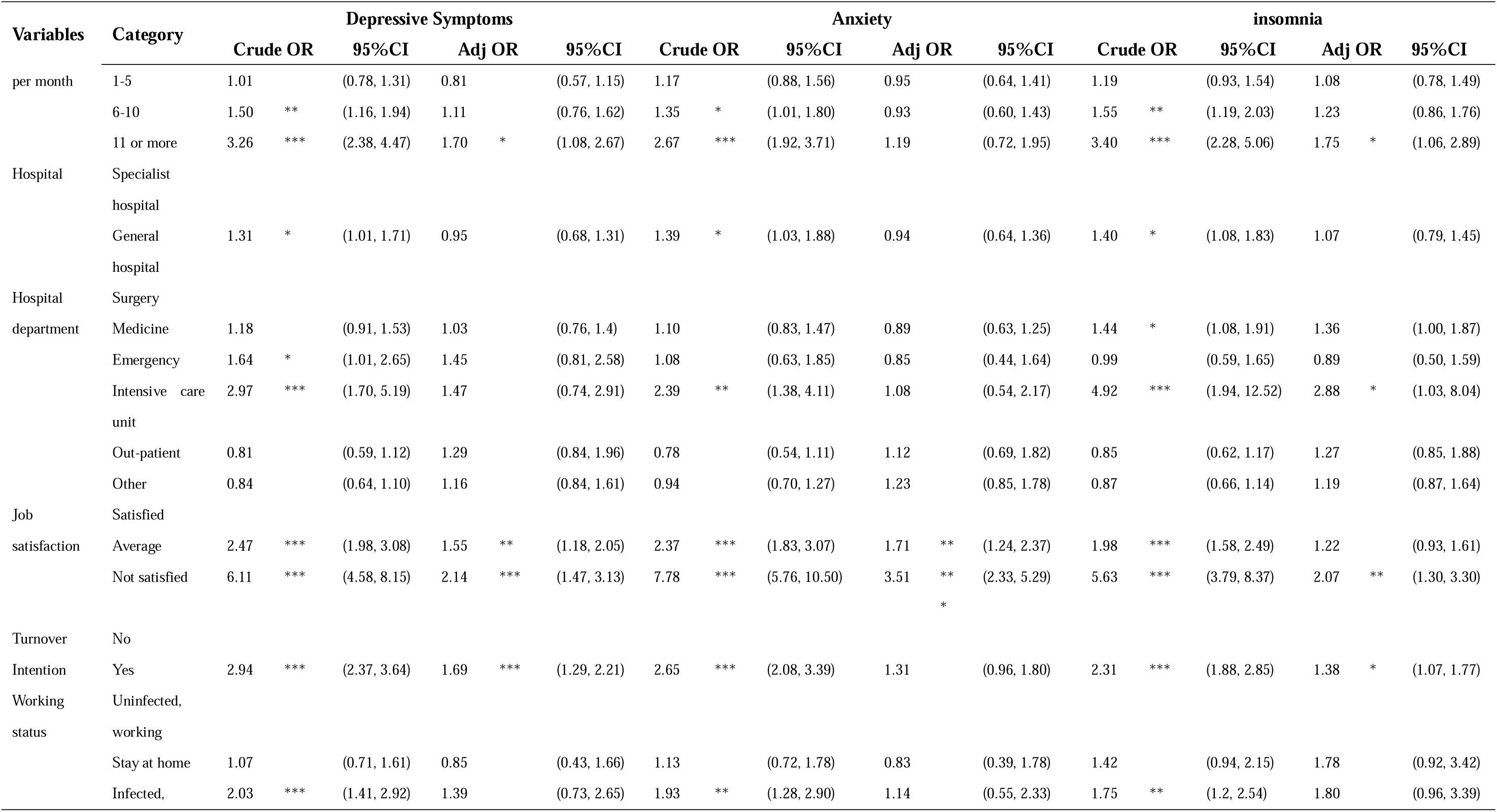
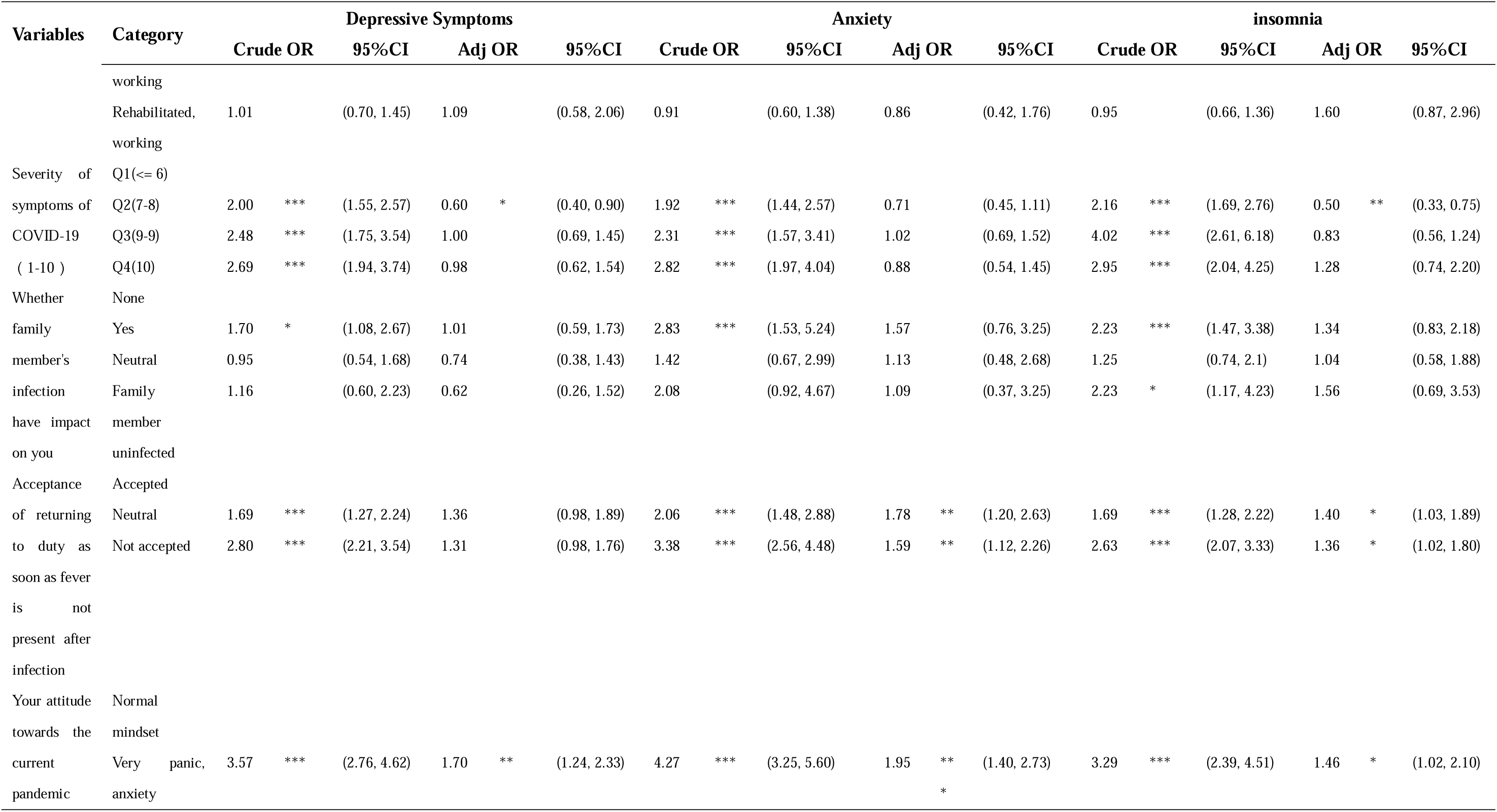
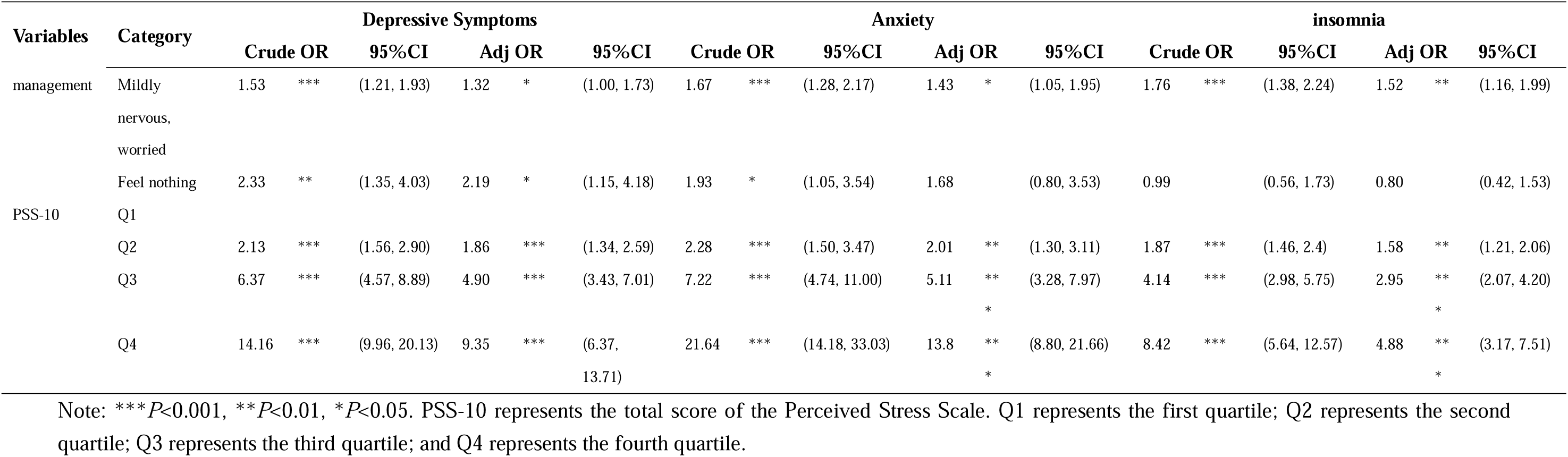
Binary logistic regression analysis of the factors associated with depressive symptoms, anxiety and insomnia.

### 4.4 Risk factors associated with anxiety symptoms

Longer working years increased the risk of anxiety. Nurses who were dissatisfied (*OR*, 3.51; 95% *CI*, 2.33-5.29) and average (*OR*, 1.71; 95% *CI*, 1.24-2.37) with their current nursing work had a higher level of anxiety than those who were satisfied. Those who had a lack of acceptance (*OR*, 1.59; 95% *CI*, 1.12-2.26) or felt neutral (*OR*, 1.78; 95% *CI*, 1.20-2.63) about returning to duty as soon as fever was not present after infection were also at greater risk of anxiety than those who accepted this. A negative attitude toward the current pandemic management and nurses with higher total PSS-10 scores also had an increased risk of anxiety (Table 3).

### 4.5 Risk factors associated with insomnia symptoms

The factor significantly associated with insomnia was turnover intention (*OR*, 1.38; 95% CI, 1.07-1.77). Nurses with 11 or more night shifts per month (*OR*, 1.75; 95% *CI*, 1.06-2.89) were at greater risk of developing insomnia than those without night shifts. The insomnia risk was higher among those working in the ICU (*OR*, 2.88; 95% *CI*, 1.03-8.04). Nurses who were dissatisfied (*OR*, 2.07; 95% *CI*, 1.30-3.30) with their current work had a higher level of insomnia than those who were satisfied. Nurses who did not accept (*OR*, 1.36; 95% *CI*, 1.02-1.80) or felt neutral (*OR*, 1.40; 95% *CI*, 1.03-1.89) about returning to duty as soon as fever was not present after infection also had a higher level of insomnia than those who accepted this. A negative attitude toward the current pandemic management and nurses with higher total PSS-10 scores also had a higher risk of insomnia (Table 3).

## 5 DISCUSSION

To the best of our knowledge, the current study represents the first attempt to examine frontline nurses’ epidemiology and correlates of psychological health during the full liberalization of COVID-19 in China using a multicenter large-scale cross-sectional design. The strengths of this survey included the multicentered sampling, the large sample size, and the use of standardized evaluation tools for depression, anxiety, perceived stress and insomnia. We conducted our research in six hospitals with large-scale treatment of COVID-19 patients, which made our sample relatively representative of frontline nurses.

In this study, it was determined that the majority of respondents were female, were aged less than 40 years, were married, had bachelor’s degrees, worked in general hospitals with a primary technical title, and had a working period between 1 and 20 years. Moreover, the present study identified certain independent risk factors associated with depressive symptoms, anxiety, and insomnia among Shenzhen frontline nurses. Our research was conducted during the critical period of full liberalization of the pandemic in China, from the COVID-19 pandemic in December 2019 to the current stage in January 2023, which was China’s first phase of comprehensive epidemic liberalization. Moreover, the study targeted participants in Shenzhen, China, which is a large population and high-income region in China. Consequently, our findings might have certain implications and reference value for frontline nurses in regions and countries currently experiencing the full liberalization of COVID-19.

Our study revealed that a significantly high proportion of frontline nurses were afflicted with depressive symptoms (69.20%), anxiety (62.51%), and insomnia (76.78%). In the survey, approximately 38.73% nurses showed moderate to severe depressive symptoms, 26.45% had moderate to severe anxiety, and 34.59% had moderate to severe insomnia. Frontline nurses revealed more severe psychological symptoms on all measurements.

Most previous studies concentrated on nurses’ psychological health at the early stage of the COVID-19 pandemic, when the psychological impacts of COVID-19 on nurses were already apparent. A cross-sectional study using the same scales as our study demonstrated that the prevalence of depression, anxiety, and insomnia among frontline health care workers in China exposed to COVID-19 was 50.4%, 44.6%, and 34.0% (Lai et al., 2020), respectively, which was lower than that in the present study. Approximately 43.61% of frontline nurses in emergency departments treating COVID-19 patients experienced depression (An et al., 2020). In research during the severe acute respiratory syndrome (SARS) outbreak, a previous infectious disease, 38.5% of nurses working in high-risk situations showed psychological symptoms (Su et al., 2007). Compared to the Singapore and India study populations during the COVID-19 outbreak, our cohort showed a higher prevalence rate of moderate to severe depressive symptoms (38.73% versus 5.30%) and anxiety (26.45% versus 8.72%) (Chew et al., 2020). However, compared to the previous Chinese study population, our research indicated a higher prevalence rate of depressive symptoms (38.73% versus 16.51%) and a slightly lower rate of anxiety (26.45% versus 28.83%) (Wang et al., 2020). Thus, public health emergencies, whether SARS or COVID-19, lead to a strong psychological impact on health care professionals, especially frontline nurses (Nie, Su, Zhang, Guan & Li, 2020). However, compared with these findings, the incidence of depressive symptoms, anxiety, and insomnia in frontline nurses in our study was particularly high. This phenomenon may be attributed to the stage of sudden full liberalization control of COVID-19, which caused a rapid increase in admissions and presentations to hospitals and consequently increased the workload of nurses. In addition, nurses working as frontline health care professionals who had direct involvement with COVID-19 patients were an independent risk factor for all psychological health symptoms (Lai., 2020). Overall, 90.83% of frontline nurses in our study were infected with COVID-19, and 33.64% had to work with COVID-19-infected patients. A previous study demonstrated that the most commonly reported physical symptoms of COVID-19 were headache (31.90%), throat pain (33.55%), anxiety (26.71%), lethargy (26.60%), and insomnia (20.97%) (Chew et al., 2020). There is a bidirectional significant association between the prevalence of physical symptoms and psychological issues, where uncomfortable physical symptoms exacerbate psychological issues and vice versa (Chew et al., 2020). Furthermore, despite being infected with COVID-19, frontline nurses still have to work, which poses a threat to their mental health. Among all health care professionals, frontline nurses have been the main force in fighting against public health crises and emergencies, and their psychological health is crucial. These results highlight the requirement for early identification and the significance of effectively assessing and treating milder or moderate symptoms before they turn into more severe and complex mental health issues.

This study revealed that frontline nurses with primary or intermediate technical titles had a higher level of depression than those with senior technical titles, which agrees with previous findings (Lai et al., 2020). A possible contributing factor was that 61.83% of frontline nurses had primary technical titles and 31.94% had intermediate titles, indicating that many nurses had fewer years of work experience (Lai et al., 2020), and particularly lacked experience caring for COVID-19 patients and critically ill patients, compared with seniors. Another possible reason may be that nurses with lower technical titles had increased odds of mental health issues and PTSD during the period of caring for the sharply increasing number of positive patients (Zheng et al., 2021). Therefore, this population should be given special attention to reduce depressive symptoms.

Our results indicated that depressive symptoms and insomnia were related to night shifts per month; frontline nurses with 11 or more night shifts per month were more likely to develop depressive symptoms and insomnia than those without night shifts. Some previous studies have demonstrated that rotating night shift work may produce circadian disruption, sleep disturbances, and health behavioral changes (Kervezee, Kosmadopoulos & Boivin, 2020), resulting in an increased risk of chronic diseases (Wu et al., 2022; Vetter et al., 2016) psychological disorders (Torquati et al., 2019), cognition impairment and mortality (Jørgensen et al., 2017). Excess night shift work is a significant health concern that may result in the deteriorating overall health of frontline nurses (Shi et al., 2022), which may explain why frequently rotating night shift work was associated with more depression and insomnia during the special period of full liberalization control of COVID-19.

This study showed that turnover intention, job satisfaction, current working status, attitude toward current pandemic management, and perceived stress scores were associated with frontline nurses’ depression, anxiety, and insomnia. During the full liberalization control of COVID-19, frontline nurses, who made enormous efforts by risking their lives in emergency departments, intensive isolation units, intensive care units, and COVID-19 patient wards, demonstrated their commitment and responsibility to their profession and their patients. Nurses who treated COVID-19-positive patients tended to be exposed to the highest risk of virus infection due to their close, frequent contact with patients and intensive work. Nurses constantly suffer from stressful situations that lead to emotional exhaustion while managing the complex care and treatment processes of COVID-19 patients. Particularly during the special period of full liberalization control of COVID-19, working for a long time in a hectic environment with a high level of stress and increasing workload caused nurses to experience burnout more rapidly (Murat, Köse & Savaşer, 2021). Moreover, the incompatibility between the ideal expectations of the nursing profession and the situations encountered in real life gives rise to turnover intention and dissatisfaction with current nursing work (Woo et al., 2020) . The long duration of uncertainty during sudden public events such as emergencies and/or disasters also affects the stress level of individuals (Murat, Köse & Savaşer, 2021).

Our study highlighted that higher stress levels were a risk factor that influenced nurses’ mental health. The acceptance of returning to duty as soon as a high fever was not present after infection was an essential risk factor associated with anxiety and insomnia. This finding was consistent with previous studies (Zheng et al., 2021). Frontline nurses constantly face a number of work-related stressors, including but not limited to heavy workloads, long work hours, meeting patients’ requirements, and irregular working schedules (Maharaj, Lees & Lal, 2018). Previous studies have noted that the ongoing stress faced by these health care professionals can have negative effects on their psychological well-being (Maharaj, Lees & Lal, 2018) , especially when they face a greater threat of public emergencies. It is widely understood that perceived stress is linked to psychological status; nurses who suffer from perceived stress are more inclined to develop depressive symptoms, anxiety, insomnia, and health-related concerns. Due to the large number of infection cases, frontline nurses worried about becoming infected through asymptomatic transmission (Chen et al., 2006), a lack of effective treatment, and stigmatization (Xiang et al., 2020). In this study, 90.83% of frontline nurses were infected with COVID-19, and 33.64% had to work while infected COVID-19. It was confirmed that COVID-19 affects all human body systems physiopathologically, especially the immune and respiratory systems, which also results in negative psychological effects (Badahdah et al., 2021). Thus, during the health crises of full liberalization control of COVID-19, nurses may have had no fever and went to work after contracting the virus, but they may have had cough, sputum, and sore throat. Nurses who experienced physical discomfort due to COVID-19 might not have sufficient time to become better adjusted to caring for affected patients and may have become more concerned about personal and family health during full liberalization control.

Furthermore, the insomnia risk was higher among frontline nurses working in the ICU. The ICU is a constantly changing, highly regulated workplace for patients in critical situations. In addition, excessive workload, shift work, resuscitation and death are proven risk factors for insomnia (Morrison & Joy, 2016). Consistent with a previous study (Lai et al., 2020), our research revealed that longer working years increased the risk of anxiety, which was possibly related to the fact that nurses with longer working experience were routinely assigned to treat and care for more patients and/or more severe patients in hospitals in Shenzhen, further increasing their anxiety. Consequently, this population should be given special attention to reduce symptom severity.

### 5.1 LIMITATIONS

This study has limitations in the generalization of the results. First, the cross-sectional nature of the study merely provided information at one time point and was unable to interpret causality. Second, the lack of follow-up data on frontline nurses’ psychological health made it difficult to understand their mental health status over time. Third, the participants completed the questionnaires using the Wenjuanxing application and mobile devices, which might result in self-selection bias. Finally, the psychological variable measures in this study were based on a self-report questionnaire. Accordingly, longitudinal studies and prospective controlled studies are suggested for future studies to provide more in-depth and useful information about the psychological health of frontline nurses, both in China and in other parts of the world.

## 6 CONCLUSION

Our findings demonstrate that the full liberalization of COVID-19 had significant psychological impacts on frontline nurses. Job satisfaction, attitude toward current pandemic management, and perceived stress were associated with depressive symptoms, anxiety, and insomnia. Furthermore, frontline nurses with 11 or more night shifts per month and turnover intentions were associated with higher depressive symptoms and insomnia. Nurses who did not accept their return to duty as soon as high fever was not present after infection were associated with higher anxiety and insomnia. Therefore, it is essential for hospitals and health care institutions to implement early and effective psychosocial support and intervention for nurses to prevent the further impact of the full liberalization of COVID-19.

## 7 IMPLIC ATIONS FOR CLINICAL PRACTICE

This is the first study to examine frontline nurses’ epidemiology and correlates of psychological health during the full liberalization of COVID-19 in China using a multicenter, large-scale, cross-sectional design. This study highlighted that frontline nurses suffered from varying degrees of depressive symptoms, anxiety, and insomnia. Early screening and preventive and promotive interventions are required to prevent a more serious psychological impact on frontline nurses.

## Data Availability

All data produced in the present study are available upon reasonable request to the authors

